# T-cell receptor sequencing identifies prior SARS-CoV-2 infection and correlates with neutralizing antibody titers and disease severity

**DOI:** 10.1101/2021.03.19.21251426

**Authors:** Rebecca Elyanow, Thomas M. Snyder, Sudeb C. Dalai, Rachel M. Gittelman, Jim Boonyaratanakornkit, Anna Wald, Stacy Selke, Mark H. Wener, Chihiro Morishima, Alex L. Greninger, Michael R. Holbrook, Ian M. Kaplan, H. Jabran Zahid, Jonathan M. Carlson, Lance Baldo, Thomas Manley, Harlan S. Robins, David M. Koelle

## Abstract

Measuring the adaptive immune response to SARS-CoV-2 can enable the assessment of past infection as well as protective immunity and the risk of reinfection. While neutralizing antibody (nAb) titers are one measure of protection, such assays are challenging to perform at a large scale and the longevity of the SARS-CoV-2 nAb response is not fully understood. Here, we apply a T-cell receptor (TCR) sequencing assay that can be performed on a small volume standard blood sample to assess the adaptive T-cell response to SARS-CoV-2 infection. Samples were collected from a cohort of 302 individuals recovered from COVID-19 up to 6 months after infection. Previously published findings in this cohort showed that two commercially available SARS-CoV-2 serologic assays correlate well with nAb testing. We demonstrate that the magnitude of the SARS-CoV-2-specific T-cell response strongly correlates with nAb titer, as well as clinical indicators of disease severity including hospitalization, fever, or difficulty breathing. While the depth and breadth of the T-cell response declines during convalescence, the T-cell signal remains well above background with high sensitivity up to at least 6 months following initial infection. Compared to serology tests detecting binding antibodies to SARS-CoV-2 spike and nucleoprotein, the overall sensitivity of the TCR-based assay across the entire cohort and all timepoints was approximately 5% greater for identifying prior SARS-CoV-2 infection. Notably, the improved performance of T-cell testing compared to serology was most apparent in recovered individuals who were not hospitalized and were sampled beyond 150 days of their initial illness, suggesting that antibody testing may have reduced sensitivity in individuals who experienced less severe COVID-19 illness and at later timepoints. Finally, T-cell testing was able to identify SARS-CoV-2 infection in 68% (55/81) of convalescent samples having nAb titers below the lower limit of detection, as well as 37% (13/35) of samples testing negative by all three antibody assays. These results demonstrate the utility of a TCR-based assay as a scalable, reliable measure of past SARS-CoV-2 infection across a spectrum of disease severity. Additionally, the TCR repertoire may be useful as a surrogate for protective immunity with additive clinical value beyond serologic or nAb testing methods.

## Introduction

Understanding the immune response to severe acute respiratory syndrome coronavirus 2 (SARS-CoV-2) is essential to inform clinical management of COVID-19 and vaccination strategies to contain the pandemic (1). Infection with SARS-CoV-2 induces both humoral and T-cell responses, but the nature and kinetics of these responses are heterogeneous, varying with disease severity and individual characteristics (2–4). Similarly, antibody titer and T-cell assays have demonstrated that SARS-CoV-2 vaccines induce humoral and/or cell-mediated immune responses, but the optimal combination of responses underlying immune correlates of protection remains undefined (5, 6). This knowledge gap is underscored by recently described viral variants that can escape antibody responses (7, 8) with relatively preserved CD4+ and CD8+ T-cell responses (9), having potential impacts on vaccine-induced immunity and the capacity for viral neutralization (10, 11).

While serologic assays are a common means of assessing prior SARS-CoV-2 infection at the population level (4, 12), it remains unclear whether the results of serologic testing correspond with long-term protective immunity (13, 14). In addition, neutralizing antibody (nAb) titers, while providing one measure of immune protection in SARS-CoV-2 infection (15), are challenging to perform, pose biohazard risks and may have limited durability over time (16). More recently, T-cell receptor (TCR) repertoire-based assays have emerged as another technology for reliable assessment of prior infection and immunity (17, 18). Such assays can be performed on standard whole blood samples of 1-2mL. Through application of SARS-CoV-2-specific classifiers leveraging thousands of public TCR sequences, that is, those sequences that are shared across individuals with a history of infection, both the presence and magnitude of the T-cell response to SARS-CoV-2 can be more fully evaluated (17). However, additional data are needed to validate the utility, performance, and advantages of these assays relative to serologic or nAb testing.

A recent study compared nAb titers with the performance of two SARS-CoV-2 IgG serology tests using blood samples collected up to 6 months after symptom onset as part of a convalescent plasma donor screening program (19). Results from this study demonstrated that these tests correlate with nAb testing, enabling better prioritization of high-titer samples for immunoglobulin donor products. Here we conduct further assessments of samples from this cohort as well as additional enrolled individuals, in order to characterize the T-cell response to SARS-CoV-2 and its relationship with antibody testing strategies and clinical indicators of disease.

## Results/Discussion

### SARS-CoV-2–specific T-cell responses are correlated with neutralizing antibody titers

TCRβ sequencing was performed on samples from 302 persons with a prior positive SARS-CoV-2 RT-PCR test in the setting of compatible illness. Of these, 55 had an additional sample from a subsequent visit with a median of 90 days between samples, for a total of 357 samples. All samples were collected during convalescence, ranging from 29-190 days after symptom onset (Supplemental Table 1). We first evaluated the association between clonal breadth and depth of the T-cell response and nAb titers. Clonal breadth was defined as the relative number of distinct SARS-CoV-2-associated T-cell clonotypes as a fraction of the overall repertoire, and clonal depth as the extent of expansion of SARS-CoV-2–associated T-cells (17). Both clonal breadth (p<1e-20, ρ = 0.54, Spearman) and depth (p<1e-20, ρ=0.46, Spearman) showed highly significant positive correlations with nAb titer (Figure 1A,B). These data suggest that, like nAb titers (15), SARS-CoV-2 T-cell response signatures may represent a potential surrogate of protective immunity arising from natural infection.

**Figure 1:**
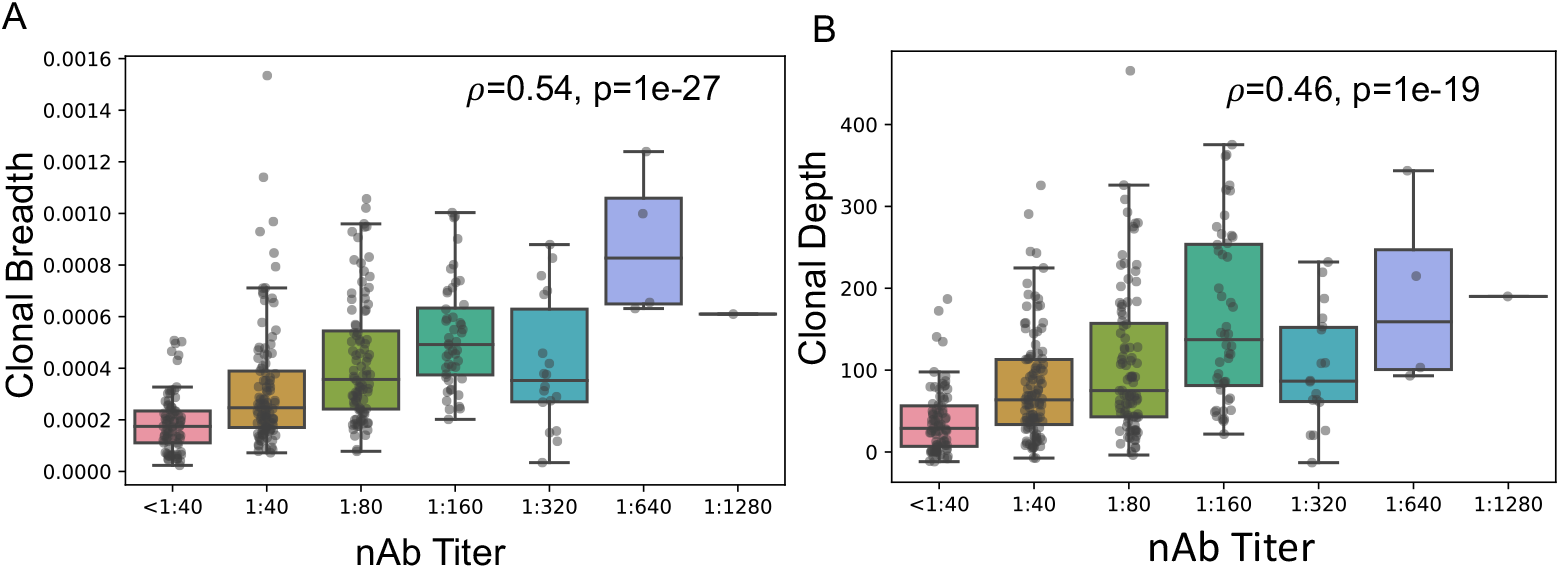
SARS-CoV-2–specific T cell responses correlate with nAb titer. Correlation of T-cell clonal breadth (A) and depth (B) with nAb titer evaluated by Spearman’s rank-order correlation.

To further characterize the correlation between the T-cell response and nAb titers, we assigned a subset of TCR sequences to specific antigens and class I or class II HLA restriction based on data from multiplexed antigen stimulation assays (17, 20). These TCRs are associated with cytotoxic (CD8+) and helper (CD4+) cellular immune responses, respectively. For helper (CD4+) T cells, we could assign 769 TCRs to antigens from SARS-CoV-2 spike protein, 362 to nucleocapsid phosphoprotein, and 474 to other viral proteins. We then evaluated correlations of nAb titers to these assigned TCR sets; partial correlations were applied to account for correlations in immune response to different antigens which may result from natural biologic variation, such as systemic immune activation or total viral burden (described in greater detail in the Supplementary Methods). These analyses revealed that the clonal breadth and depth of class II-associated TCRs for spike protein and nucleocapsid phosphoprotein remain correlated with nAb titer after partial correlation, while TCRs for the remaining antigens do not (Supplemental Figure 2A-C). Of interest, TCRs assigned to CD8+ T cells were not correlated to nAb titers, suggesting that the primary origin of the nAb correlation with overall SARS-CoV-2-specific TCR breadth and depth is the helper T-cell response. While most nAbs are presumed to be targeting the spike protein (15), these results suggest that CD4+ cells responding to antigens from spike as well as some other proteins may support the development of functional humoral immunity.

### SARS-CoV-2–specific T-cell responses are correlated with clinical measures of COVID-19 severity

Previous analyses involving this cohort revealed significant associations between nAb response and a number of important clinical correlates, including older age, male sex, the presence of fever, difficulty breathing, and hospitalization denoting more severe disease (19). We observed that the clonal breadth of the SARS-CoV-2 T-cell response is significantly correlated (p< 0.05) with each of these variables by both single and multi-variable regression (Figure 2). Clonal depth was also significantly correlated (p<0.05) with each variable except difficulty breathing (Supplemental Figure 1). These associations are independent of the number of unique T-cell rearrangements, which was included in the multi-variable regression. The association between magnitude of the T-cell response and clinical indicators of disease severity is consistent with other reports showing higher T-cell responses in symptomatic versus asymptomatic individuals that can persist months after infection (3, 21). We hypothesize that increased viral load, longer viral persistence, and/or higher levels of immune activation during acute SARS-CoV-2 infection may in part underlie the association between more severe COVID-19 illness and greater recruitment and long-term durability of the T-cell response. T cell breadth and depth was also higher in men and older adults (Figure 2D,E; Supplemental Figure S1D,E), consistent with increased rates of severe illness and hospitalization seen in these groups (22).

**Figure 2:**
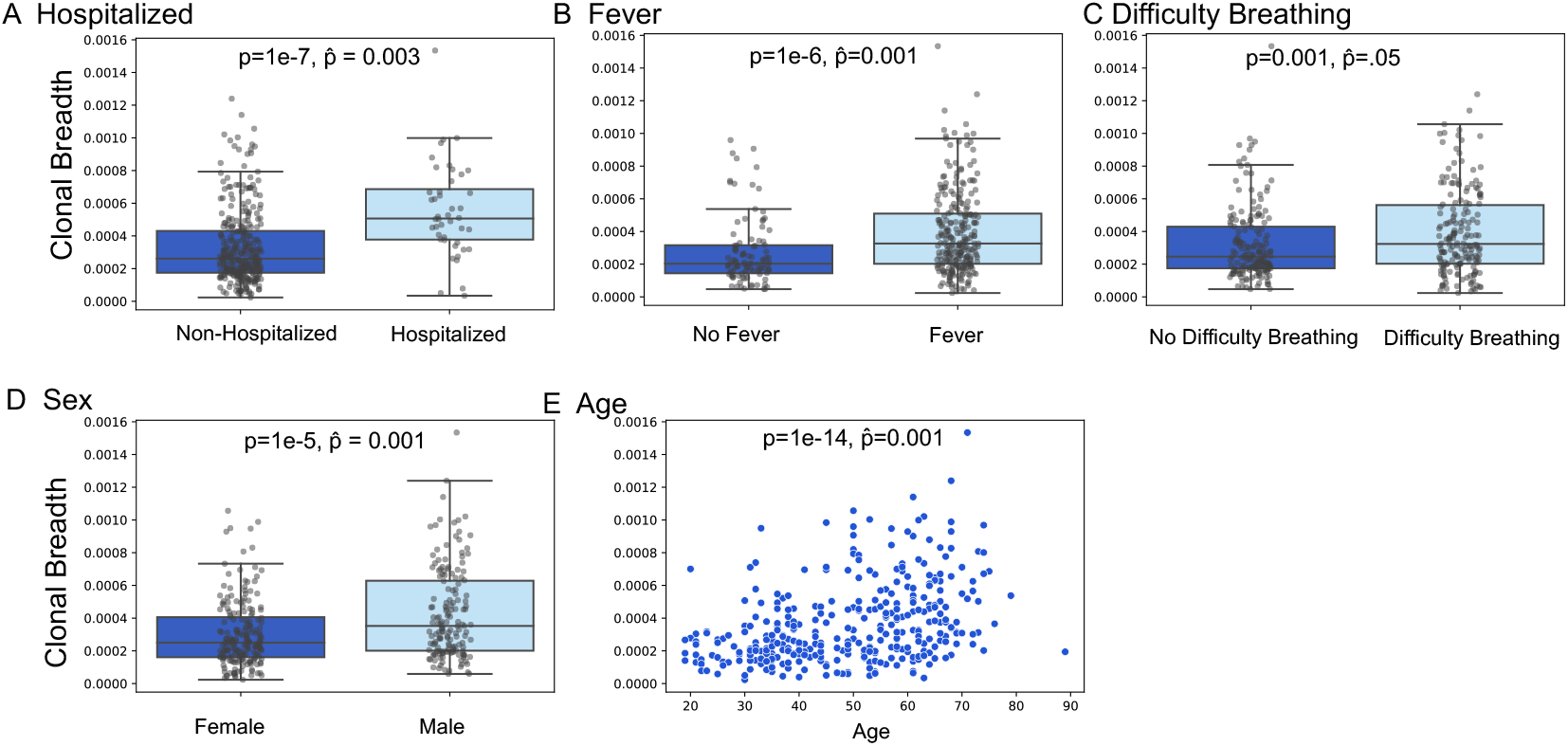
Association of T-cell clonal breadth with clinical variables. Association of clonal breadth with (A) hospitalization, (B) fever, (C) difficulty breathing, (D) sex, and (E) age. *P* values indicated by p and 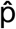 are for univariate Mann-Whitney U test and multivariable linear regression (with variables age, sex, hospitalization, fever, difficulty breathing, and TCR rearrangements) respectively.

### SARS-CoV-2-specific T-cell responses have higher diagnostic sensitivity than serology in identifying past infection, particularly in non-hospitalized cases

Based on identification of public SARS-CoV-2–specific T-cell signatures shared across individuals, a classifier was developed to diagnose recent and past SARS-CoV-2 infection (17) in previously-confirmed RT-PCR-positive cases that has been validated in several independent data sets (18, 21). Optimization and application of this TCR classifier (further described in Methods) as a test for past SARS-CoV-2 infection yielded a sensitivity of 88.8% across all samples and timepoints, with a specificity of 99.8% in a control set of 1,657 pre-pandemic samples sequenced prior to 2020 (Figure 3A; Supplemental Table 2). Consistent with the observation that TCR repertoire breadth and depth are greater in individuals with more severe disease (Figure 2, Supplemental Figure 1), we found that sensitivity of the classifier was higher in hospitalized (93.2%) compared to non-hospitalized cases (88.2%; Supplementary Table 2). Although the classifier (log-odds) scores decreased slightly with time from symptom onset (ρ = −0.15, p=0.01, Spearman), the assay maintained a sensitivity of 95% for samples collected >150 days after symptom onset (Figure 3B). Together, these results support the utility of the TCR-based assay as a sensitive measure of prior disease.

**Figure 3:**
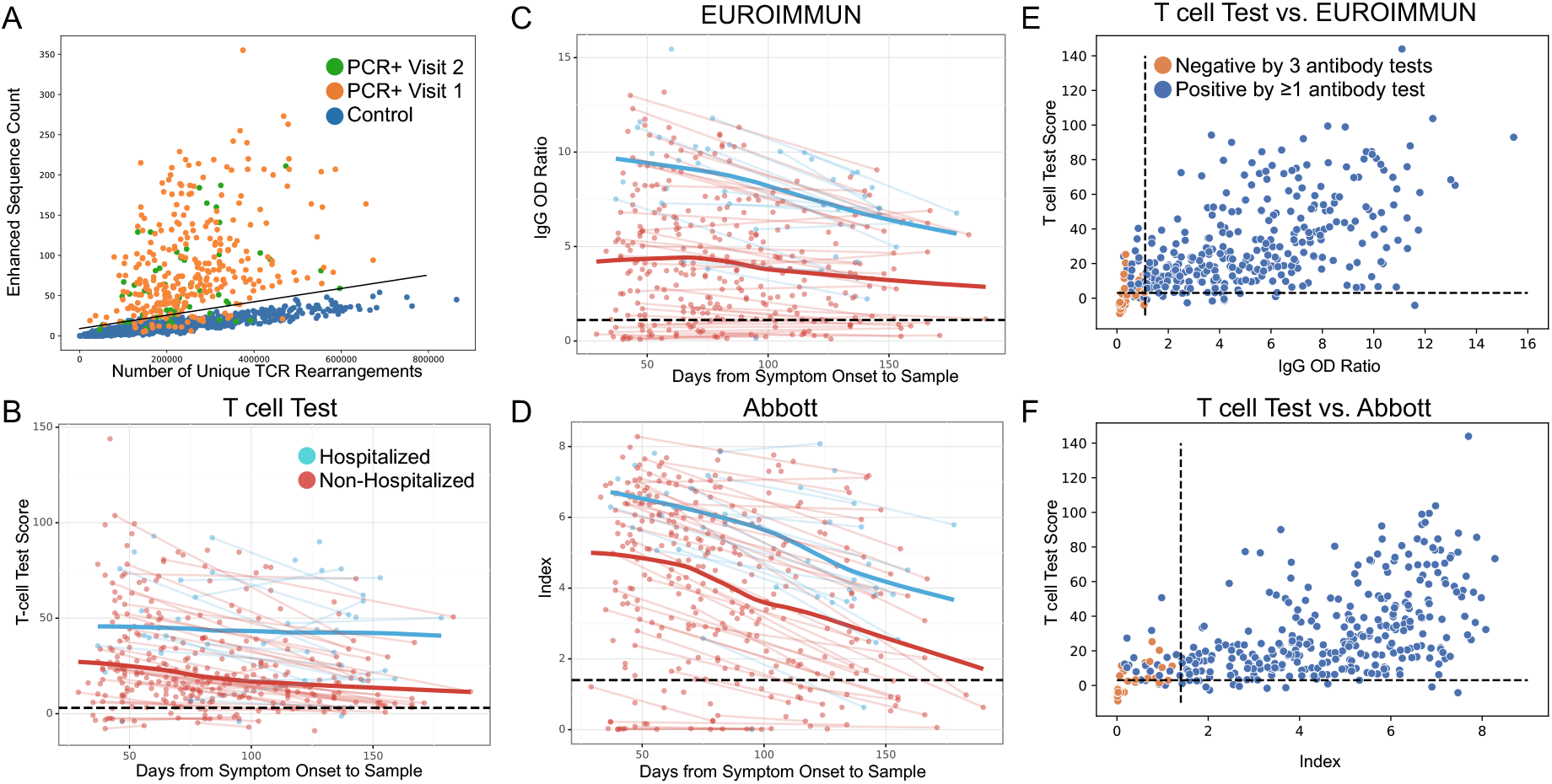
Comparison of TCR-based assay with serological assays. (A) Number of enhanced sequences associated with SARS-CoV-2 infection for RT-PCR–confirmed samples collected at visit 1 (orange) or visit 2 (green). The axes are the two parameters comprising the T-cell Test: enhanced sequence count out of 4,287 enhanced sequences (y-axis) and number of unique TCR rearrangements (x-axis). Blue dots represent 1,657 control samples collected before January 2020. All samples were held out from training the classifier. (B) T-cell Test score, (C) EUROIMMUN IgG OD ratio, and (D) Abbott index as a function of days from symptom onset to sample, indicated as hospitalized (blue) or non-hospitalized (red) individuals, with trend lines connecting visit 1 and visit 2 sample points from the same subject. Blue and red bold trend lines indicate smoothed mean (locally estimated scatterplot smoothing = LOESS [Cleveland 1981]) for hospitalized and non-hospitalized individuals, respectively. (E) T-cell Test scores in RT-PCR-confirmed samples compared to EUROIMMUN IgG OD ratio and (F) Abbott index. Samples classified negative by all three antibody tests (EUROIMMUN, Abbott, and nAb titer) are highlighted in orange. Black dashed lines indicate cutoffs for positivity/negativity. The cutoff used for positivity for nAb is 1:40 recognizing that some samples could have neutralizing titers of < 1:40.

Next, we compared results from the TCR-based assay with two serology assays which were interpreted using manufacturer-suggested cutoffs (1.1 for EUROIMMUN Anti-SARS-CoV-2 ELISA, 1.4 for Abbott Architect; www.fda.gov). The EUROIMMUN assay measures IgG binding to the S1 domain of the SARS-CoV-2 spike protein, and the Abbott Architect assay measures IgG binding to the SARS-CoV-2 nucleocapsid protein. Across the entire cohort and inclusive of all timepoints assessed, these assays had sensitivities significantly lower than TCR-based testing (p=0.01, mid-p McNemar’s test; Table 1, Supplemental Table 2) (19). Notably, when performance was assessed at specific sampling timepoints in the months following initial illness, both serology assays showed steeper declines in quantitative diagnostic scores over time relative to the TCR-based assay (Figure 3C,D), with the greatest differences in sensitivity observed beyond 5 months (>150 days) from initial symptoms (p<0.03 comparing the TCR-based assay to EUROIMMUN and Abbott, mid-p McNemar’s test; Table 1, Supplemental Table 2).

**Table 1:** Comparison of sensitivities for T-cell Test and commercial serological assays.

| Days from Symptom Onset to Sample | T cell Test | EUROIMMUN (anti-S1 IgG) | Abbott (anti-NP IgG) |
| --- | --- | --- | --- |
| All Data | 317/357 (88.8%) | 297/357 (83.2%) | 300/357 (84.0%) |
| > 100 Days | 103/117 (88.0%) | 95/117 (81.2%) | 90/117 (76.9%) |
| > 150 Days | 19/20 (95.0%) | 14/20 (70.0%) | 10/20 (50.0%) |
| All Non-Hospitalized | 276/313 (88.2%) | 254/313 (81.2%) | 257/313 (82.4%) |
| Non-Hospitalized > 100 Days | 81/94 (86.2%) | 72/94 (76.6%) | 68/94 (72.3%) |
| Non-Hospitalized > 150 Days | 15/16 (93.8%) | 10/16 (62.5%) | 6/16 (37.5%) |

We then explored the performance of TCR and serology tests among hospitalized and non-hospitalized individuals (Supplemental Table 3). All 3 tests exhibited high sensitivity (93.2% or higher) for identifying prior SARS-CoV-2 infection in hospitalized individuals, with no significant difference in performance between tests (p=0.15, mid-p McNemar’s test) and consistently high sensitivity >100 days after symptom onset (Supplemental Table 3). However, for non-hospitalized individuals, the sensitivity of the TCR-based test was significantly higher than serology tests (p=0.01, mid-p McNemar’s test; Table 1). Notably, in non-hospitalized individuals, wider differences in sensitivity were seen for samples tested >100 days from symptom onset (86.2%, 76.6%, and 72.3% for the T-cell test, EUROIMMUN, and Abbott, respectively; p=0.01, mid-p McNemar test; Table 1). Taken together, these findings suggest that the rate of signal decline over time may be faster for antibody-based tests relative to T-cell testing, particularly in non-hospitalized individuals (Figure 3C,D), consistent with data indicating that some individuals may undergo loss of detectable antibodies or seroreversion (23).

### SARS-CoV-2-specific T-cell responses are detectable in a large proportion of convalescent cases testing negative by nAb or serology

Figure 3 (E,F) and Supplemental Figure 3A show discordant results from antibody testing compared to T-cell testing on all samples. While the EUROIMMUN, Abbott, and nAb tests identified some samples as SARS-CoV-2-positive that were classified as negative by T-cell testing, this was offset by a greater number of samples identified as SARS-CoV-2-positive that were not identified by each of the 3 antibody assays, resulting in 24, 21, and 41 more samples being correctly classified by T-cell testing compared to EUROIMMUN, Abbott, and nAb respectively. Notably, T-cell testing was able to identify 68% (55/81) of convalescent SARS-CoV-2 samples with nAb titers below the lower limit of detection (1:40); nearly all (53/55) of these samples originated from non-hospitalized cases. Additionally, T-cell testing identified 37% (13/35) of samples which were classified as negative by all three antibody assays, all originating from non-hospitalized individuals (Figure 3E,F; Supplemental Figure 3B). Notably, the 22 samples without detectable immune responses on any test originated from patients with mild disease, with a lower reported incidence of fever compared to other samples (p=0.01, T-test) and only one patient with hospitalization. These results are consistent with other published reports describing that some individuals generate a T-cell response to SARS-CoV-2 infection without detectable antibodies, particularly in mild disease (23– 25).

Results from this study provide evidence for robust and persistent T-cell responses to SARS-CoV-2 infection, particularly in patients with more severe disease who required hospitalization. The depth and breadth of T-cell responses correlate strongly with nAb titers, supporting the potential role for TCR repertoire sequencing as a surrogate for SARS-CoV-2 protective immunity that reflects both T-cell and humoral compartments. Finally, we observe that T-cell testing can identify SARS-CoV-2 responses in high proportions of individuals who do not demonstrate antibody responses on either nAb or serologic assays, suggesting that such assays may provide incomplete information on immune protection, particularly in individuals with less severe illness. This observation underscores that measuring both cellular (via CD4+ and CD8+ T cells) and humoral immunity is likely necessary for thorough assessment of the immune response to SARS-CoV-2 infection, and that T-cell testing provides important additive diagnostic value. This is especially relevant for SARS-CoV-2 vaccination strategies, where recently updated FDA guidance recommends evaluation of vaccine immunogenicity endpoints via nAb response, as well as exploration of the potential impact of emerging viral variants on vaccine-induced immunity (26). From our own experiments mapping TCRs to specific viral antigens (17) as well as other reports (9), we expect that the majority of T-cell responses will not be impacted by viral strain variations; we are conducting additional research to directly assess this in clinical samples.

We also show that a SARS-CoV-2-specific TCR-based test has high sensitivity for diagnosis of prior SARS-CoV-2 infection in individuals up to at least 6 months following initial infection. We have previously described the clinical validation and performance of a TCR-based assay (T-Detect) for diagnosing past SARS-CoV-2 infection with high sensitivity, ∼100% specificity, equivalent or higher sensitivity compared to commercial serologic testing, and lack of pathogen cross-reactivity (18). In the present study, using an updated classifier that includes additional filtering of the TCR sequence list (described in Methods), we observed that performance of the TCR-based test was equivalent to serology testing in hospitalized individuals and was more sensitive than serology in non-hospitalized individuals, particularly at timepoints beyond 100 days from initial onset of symptoms. The largest difference in performance was observed at the latest time points (>150 days), but we note that the sample size was limited to 20 samples over these ranges and additional research is needed to confirm the long-term performance. As only a minority of individuals with COVID-19 require hospitalization (5.3%, derived from a CDC data summary of US cases) (27), the improved performance observed in symptomatic but non-hospitalized individuals supports the utility of T-cell based testing compared to serology for monitoring past infection in the real-world where the vast majority of infections occur in the outpatient setting.

We acknowledge several limitations in the present study, including smaller sample sizes at later timepoints analyzed, restriction of the cohort to symptomatic cases only, and limited ethnic diversity of the sample population. Nevertheless, these results collectively indicate that analysis of the TCR repertoire from small-volume standard blood samples may be a useful surrogate for evaluating past infection and immune protection months after COVID-19 illness that could overcome several challenges in nAb and serologic testing, including labor intensity, biohazard risks, scalability, incomplete or absent antibody signal in non-severe illness, or limited antibody durability over time. Future studies across diverse populations and longer-term follow-up are needed to better define the nature and duration of the detectable T-cell response and its utility as a biomarker for assessing both natural and vaccine-mediated immunity.

## Data Availability

T-cell repertoire profiles and antigen annotation data from the multiplexed antigen-stimulation experiments will be available as part of the ImmuneCODE data resource (Nolan 2020), and can be downloaded from the Adaptive Biotechnologies immuneACCESS site under the immuneACCESS Terms of Use at https://clients.adaptivebiotech.com/pub/covid-2020.

## Acknowledgements

The authors wish to acknowledge Kristin MacIntosh, Danniel Zamora, Sarah McGuffin, Adrienne E. Shapiro, Victoria L. Campbell, Christopher L. McClurkan, Lichen Jing, Robin Gross, Janie Liang, Elena Postnikova, Steven Mazur, Vladimir V. Lukin, Anu Chaudhary, Marie K. Das, Susan L. Fink, Andrew Bryan, Keith R. Jerome, and Terry B. Gernsheimer. The authors also acknowledge the staff of the University of Washington Virology Research Clinic, and the volunteer subjects.

## Author Contributions

D.M.K. and A.W. designed the initial study from which these samples were drawn. This follow-up investigation was conceived by D.M.K. and A.W. with I.M.K., T.M.S., S.C.D., L.B. H.S.R.. Data analysis of T-cell repertoires was performed by R.E., T.M.S., R.M.G. with support from J.B., A.W., S.S, M.H.W., C.H., A.L.G., M.R.H., D.M.K. on prior clinical and antibody data. R.E., T.M.S., S.D., R.M.G., L.B., T.M., H.S.R, D.M.K. contributed to the interpretation of the data. R.E., T.M.S., S.C.D., R.M.G., D.M.K. wrote the manuscript, with additional contributions and edits by J.B., A.W., S.S., M.H.W., C.M., A.G., M.R.H., I.M.K., H.J.Z., J.M.C., L.B., T.M., and H.S.R..

## Author Declarations

R.E., T.M.S., R.M.G., and I.M.K. declare employment and equity ownership with Adaptive Biotechnologies. S.C.D. declares employment and equity ownership with Adaptive Biotechnologies and employment with Stanford University School of Medicine. A.W. declares institutional support from NIH. A.G. declares consulting fees with Abbott; institutional support from Abbott, Merck, and Gilead; and family employment at Labcorp. H.J.Z. and J.M.C. declare employment and equity ownership with Microsoft Corporation. T.M., L.B., and H.S.R. declare leadership, employment, and equity ownership with Adaptive Biotechnologies. D.M.K. declares institutional support from NIAID Contract 75N93019C00063. J.B., S.S., M.H.W., C.M., and M.R.H. declare no conflicts of interest.

## Funding

Supported by Adaptive Biotechnologies. The project was funded in part by the Frederick National Laboratory for Cancer Research with support from the National Institute for Allergy and Infectious Diseases (NIAID) under contract 75N91019D00024. This project has been funded in part with federal funds from the National Institute of Allergy and Infectious Diseases (NIAID), National Institutes of Health (NIH), U.S. Department of Health and Human Services (DHHS), under contract HHSN272201800013C. M.R.H. performed this work as an employee of Laulima Government Solutions, LLC. This work was also supported by NIH contract 75N93019C0063 (D.M.K.). The content of this publication does not necessarily reflect the views or policies of the DHHS or of the institutions and companies with which the authors are affiliated. This work was also supported by the Fred Hutchinson Joel Meyers Endowment (J.B.), Fast-Grants (J.B.), a new investigator award from the American Society for Transplantation and Cell Therapy (J.B.). Editorial support was provided by Melanie Styers of BluPrint Oncology Concepts, funded by Adaptive Biotechnologies.

## Supplemental Materials

### Methods

#### Clinical sample collection and serology assays

The Virology Research Clinic at the University of Washington began enrollment in an IRB-approved study in April 2020, and all participants provided informed written consent. The study recruited persons with a laboratory-confirmed SARS-CoV-2 infection who volunteered to be considered for convalescent plasma donation. The characteristics of the initial 250 persons in the cohort have been recently published (Boonyaratanakornkit 2021). The methods used for serologic assays for antibody responses were included in that report. We continued to recruit subjects and the present report extends this cohort to 302 persons.

#### Immunosequencing of T-cell receptor (TCR) repertoires

Genomic DNA was extracted from cryopreserved peripheral blood mononuclear cells. As much as 18 mg of input DNA was then used to perform immunosequencing of the CDR3 regions of TCRβ chains using the ImmunoSEQ assay. Briefly, input DNA was amplified in a bias-controlled multiplex PCR, followed by high-throughput sequencing. Sequences were collapsed and filtered to identify and quantitate the absolute abundance of each unique TCRβ CDR3 region amino acid sequence for further analysis as previously described (Robins 2009; Robins 2012; Carlson 2013).

#### T-Detect model to characterize the T-cell response to SARS-CoV-2

Due to the extreme diversity of CDR3 sequences, a majority of TCRs are private to any given individual. However, a subset of TCRs can be detected in multiple individuals and may be the result of exposure to a common antigen (Robins 2010). These public TCRs serve as a biomarker of disease and have been used to build diagnostic classifiers for CMV exposure/serostatus (Emerson 2017), and more recently for SARS-CoV-2 (Snyder 2020). In this work, classification of prior SARS-CoV-2 infection, as well as the clonal depth and breadth of the T-cell response, were determined using methods based published by Snyder et al. (Snyder 2020). Briefly, TCR repertoires from 784 unique cases of RT-PCR confirmed SARS-CoV-2 infection and 2,447 healthy controls collected before 2020 were compared using one-tailed Fisher’s exact tests to identify 4,469 public TCRβ amino acid sequences (“enhanced sequences”) significantly enriched in SARS-CoV-2–positive samples. None of the samples used for model training were from the clinical cohort investigated in the present study.

Following initial selection of enhanced sequences, some filtering of the sequence list was performed to remove potential false positives. Specifically, we reasoned that TCRs associated with CMV seropositivity or HLA alleles in non-COVID-19 healthy populations were unlikely to be specific to SARS-CoV-2. We therefore identified TCRs associated with CMV seropositivity or any of over a hundred HLA-I or HLA-II subtypes using a one-tailed Fisher’s exact test applied to TCRβ repertoires of ∼2,000 healthy controls with available HLA genotyping and CMV serotyping data. A total of 182 candidate SARS-CoV-2-associated TCR sequences were also associated with an HLA subtype or CMV seropositivity and were removed, leaving 4,287 enhanced sequences.

The final list of enhanced sequences was used to develop a classifier predicting recent or past infection with SARS-CoV-2 using a simple two-feature logistic regression, with independent variables E and N, where E is the number of unique TCRβ DNA sequences that encode an enhanced sequence and N is the total number of unique productive TCRβ DNA sequences in that subject. We define the T cell test score to be the log-odds of the probability of this logistic regression model. A decision boundary on this T cell test score representing 99.8% specificity on 1,657 controls was used to define the test-positive threshold used in the present study; this model was identical to that used in another recent analysis (Gittelman 2021).

The narrowed list of enhanced sequences was also used to calculate the clonal depth and breadth using the same formulae described in more detail by Snyder, et al. (Snyder 2020). Briefly, treating unique TCR DNA sequences observed in a repertoire as distinct clonal lineages, clonal breadth represents the fraction of all observed lineages in a repertoire that are lineages associated to SARS-CoV-2. Clonal depth accounts for the extent of clonal expansion of each lineage. With ti representing the total number of T cells observed for lineage i, N representing the total number of T cells in a sequenced repertoire, and D representing the set of all disease-associated lineages, we estimate the clonal generations for each lineage as log_2_(1+*t*_*i*_). Clonal depth, normalizing for depth of sampling, is calculated as Σ_*i*∈𝒟_log_2_(1+*t*_*i*_)−log_2_(*N*).

#### Estimating the SARS-CoV-2 protein antigen-specific T-cell response

Public TCRs were assigned to SARS-CoV-2 antigens by cross-referencing enhanced sequences identified via our case/control design with TCRs observed in multiplexed antigen-stimulation experiments, both described in prior work (Snyder 2020). To maximize the number of TCR antigen assignments, we identified a set of public TCRs from an augmented sample of repertoire data comprising prior training and validation repertoires, 1,143 additional SARS-CoV-2-positive samples, and over 1,800 samples identified as SARS-CoV-2-negative from another large study (Gittelman 2020). The final sample of repertoires, consisting of 1,927 cases and 4,135 controls, was used to identify ∼500,000 candidate public SARS-CoV-2-specific TCRs with a Fisher’s Exact Test *P* value < 0.05. We cross-referenced this list of TCRs with a set of ∼400,000 TCRs independently derived from multiplexed antigen-stimulation experiments to yield 3,381 overlapping TCRs in both datasets with protein and CD4+/CD8+ assignments determined based on antigen-stimulation experiments.

The Pingouin package in Python (Vallat 2018) was used to calculate Spearman rank correlations between antibody titers and CD4+ and CD8+ T-cell responses and report the two-sided significance. To disentangle confounding correlations, partial Spearman rank correlations were calculated between spike, nucleocapsid phosphoprotein, and other antigen specific T-cell responses and antibody titers; two-sided significance was reported. Partial correlation coefficients and *P* values are denoted with tildes when included in figures. When calculating partial correlations between antibody titers and specific categories of T-cell responses (CD4+ T-cell response specific to spike, nucleocapsid phosphoprotein, and all other assayed proteins), the other T-cell groups served as covariates. These partial correlations describe the correlation between two variables that cannot be explained by the covariates, representing a conservative measure of correlation.

---

**Supplemental Table 1:**
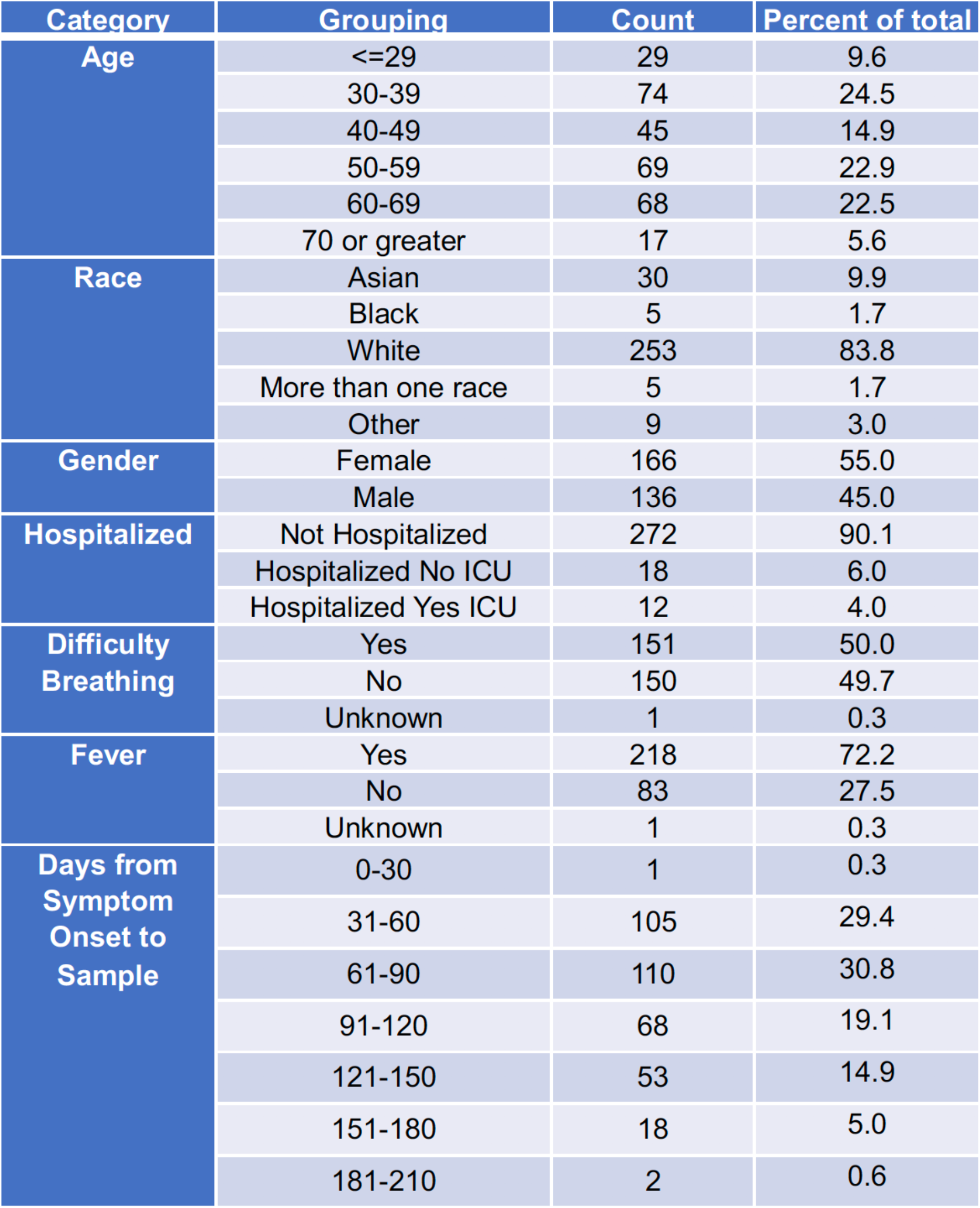
Sample demographics.

**Supplemental Table 2:** **Longitudinal sensitivity of the T-Cell Test, EUROIMMUN, and Abbott tests by time from symptom onset (30-day bins)**.

| Days from Symptom Onset to Sample | T cell Test |  | EUROIMMUN (anti-S1 IgG) |  | Abbott (anti-NP IgG) |  |
| --- | --- | --- | --- | --- | --- | --- |
|  | Sensitivity | 95% CI | Sensitivity | 95% CI | Sensitivity | 95% CI |
| All Data | 317/357 (88.8%) | 85.1-91.9 | 297/357 (83.2%) | 79.0-86.8 | 300/357 (84.0%) | 79.8-87.7 |
| 29-50 | 60/70 (85.7%) | 77.1-92.9 | 57/70 (81.4%) | 71.4-90.0 | 59/70 (84.3%) | 75.7-91.4 |
| 51-100 | 154/170 (90.6%) | 85.9-94.7 | 145/170 (85.3%) | 80.0-90.6 | 146/170 (88.8%) | 83.5-93.5 |
| 101-150 | 84/97 (86.6%) | 79.4-92.8 | 81/97 (83.5%) | 76.3-90.7 | 80/97 (82.5%) | 75.3-89.7 |
| 150-190 | 19/20 (95%) | 85.0-100 | 14/20 (70%) | 50.0-90.0 | 10/20 (50%) | 30.0-70.0 |

**Supplemental Table 3.** **Longitudinal sensitivity of the T-cell Test, EUROIMMUN, and Abbott tests for (A) hospitalized and (B) non-hospitalized patients by 30-day bins**.

| Days from Symptom Onset to Sample | T cell Test |  | EUROIMMUN (anti-S1 IgG) |  | Abbott (anti-NP IgG) |  |
| --- | --- | --- | --- | --- | --- | --- |
|  | Hospitalized | Non-Hospitalized | Hospitalized | Non-Hospitalized | Hospitalized | Non-Hospitalized |
| All Data | 41/44 (93.2%) | 276/313 (88.2%) | 44/44 (97.7%) | 254/313 (81.2%) | 42/45 (95.5%) | 257/313 (82.4%) |
| 29-50 | 3/4 (75%) | 57/66 (86.4%) | 4/4 (100%) | 53/66 (80.3%) | 4/4 (100%) | 55/66 (83.3%) |
| 51-100 | 16/17 (94.1%) | 138/153 (90.2%) | 16/17 (94.1%) | 129/153 (84.3%) | 16/17 (94.1%) | 135/153 (88.2%) |
| 101-150 | 18/19 (94.7%) | 66/78 (84.6%) | 19/19 (100%) | 62/78 (79.5%) | 18/19 (94.7%) | 62/78 (79.5%) |
| 150-190 | 4/4 (100%) | 15/16 (93.8%) | 4/4 (100%) | 10/16 (62.5%) | 4/4 (100%) | 6/16 (37.5%) |

**Supplemental Figure 1:**
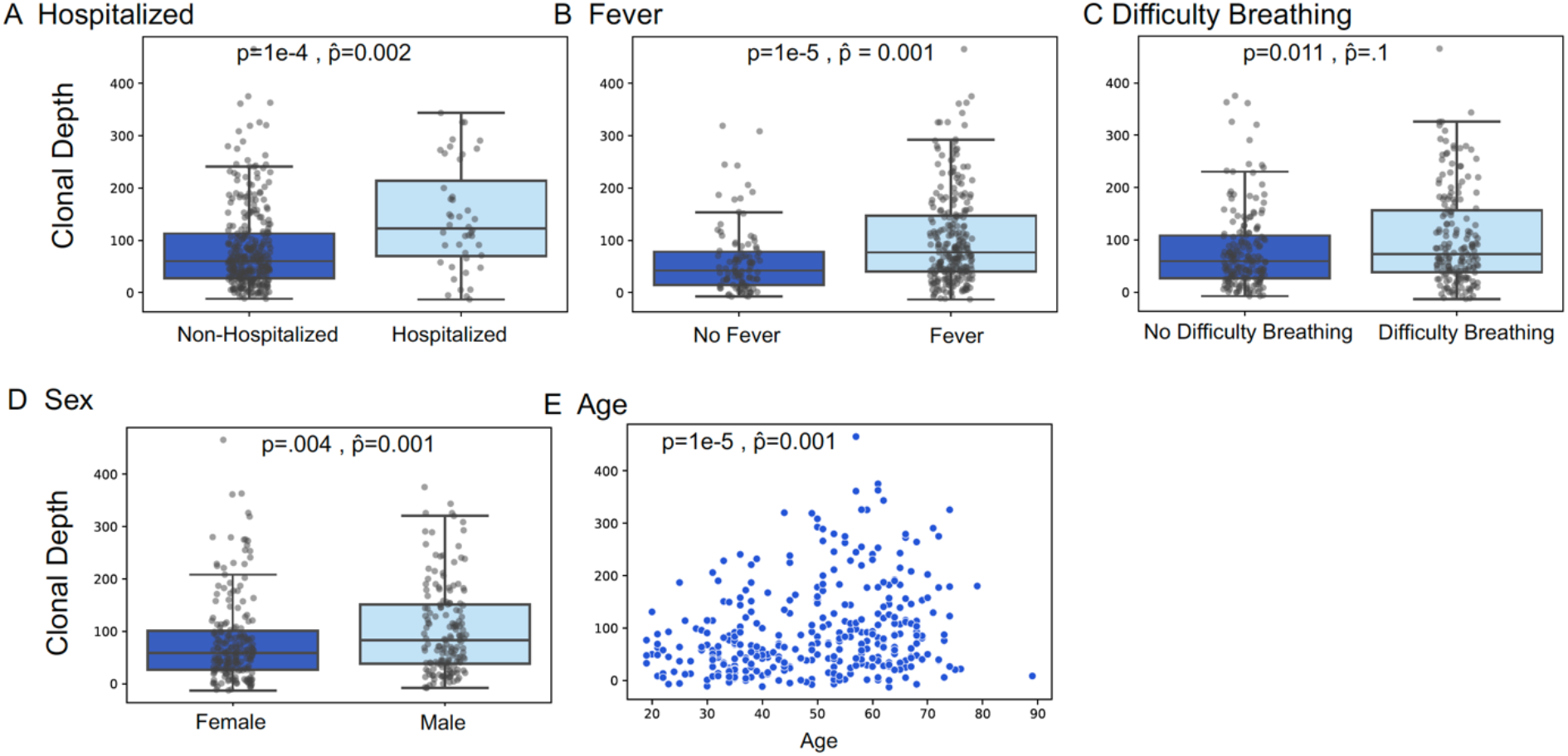
Association of T-cell clonal depth with clinical variables. Association of T-cell clonal depth with (A) hospitalization, (B) fever, (C) difficulty breathing, (D) sex, and (E) age. *P* values indicated by p and 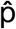 are for univariate Mann-Whitney U test and Multivariate Linear Regression respectively.

**Supplemental Figure 2:**
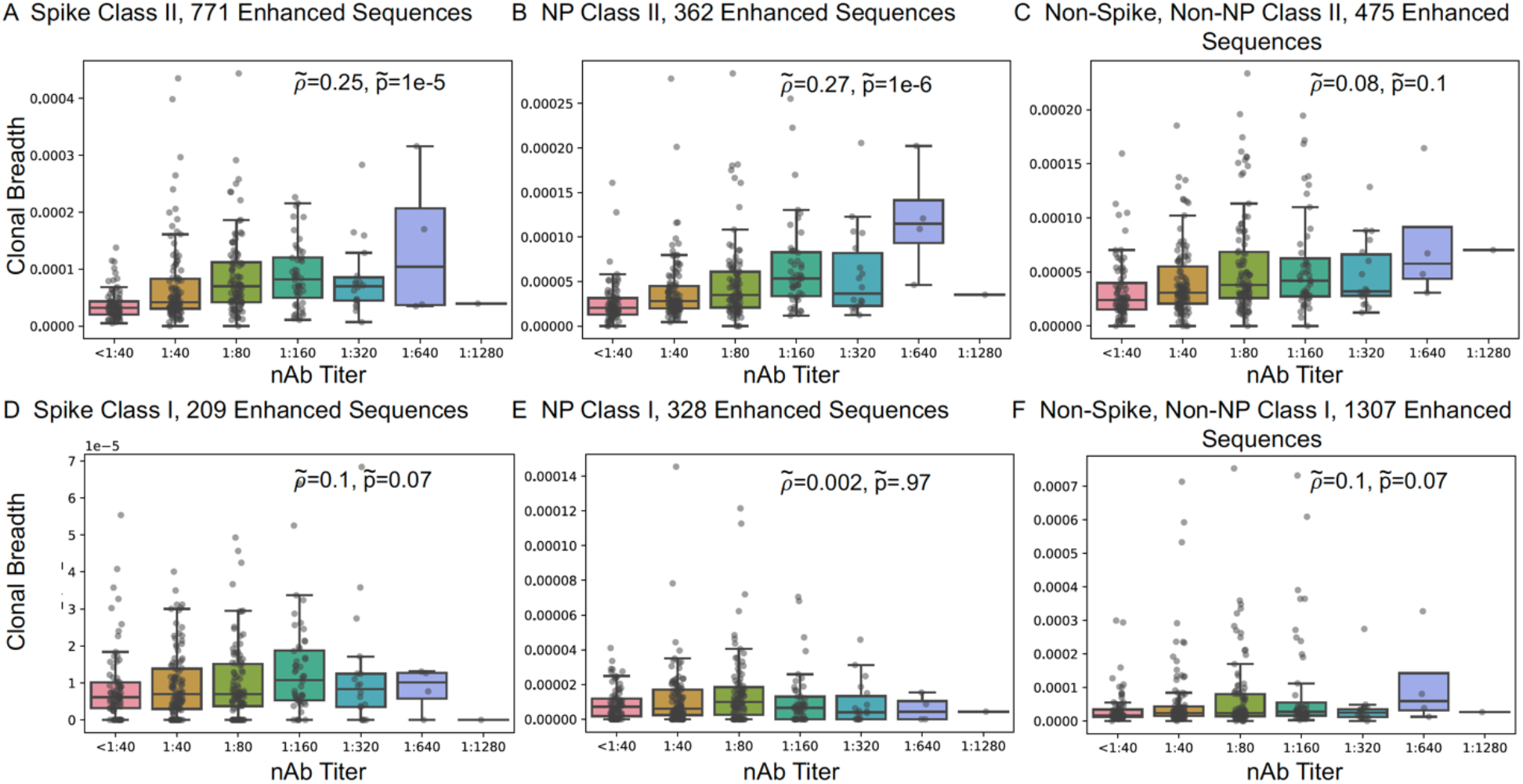
Correlation of clonal breadth of spike class II-associated T-cells. (A), nucleocapsid protein (NP) class II-associated T-cells (B), other class II T-cells (C) with nAb titer, spike class I-associated T-cells (D), nucleocapsid protein (NP) class I-associated T-cells (E), and other class I T-cells (F) with nAb titer. *P* values for partial correlation indicated by 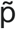.

**Supplemental Figure 3:**
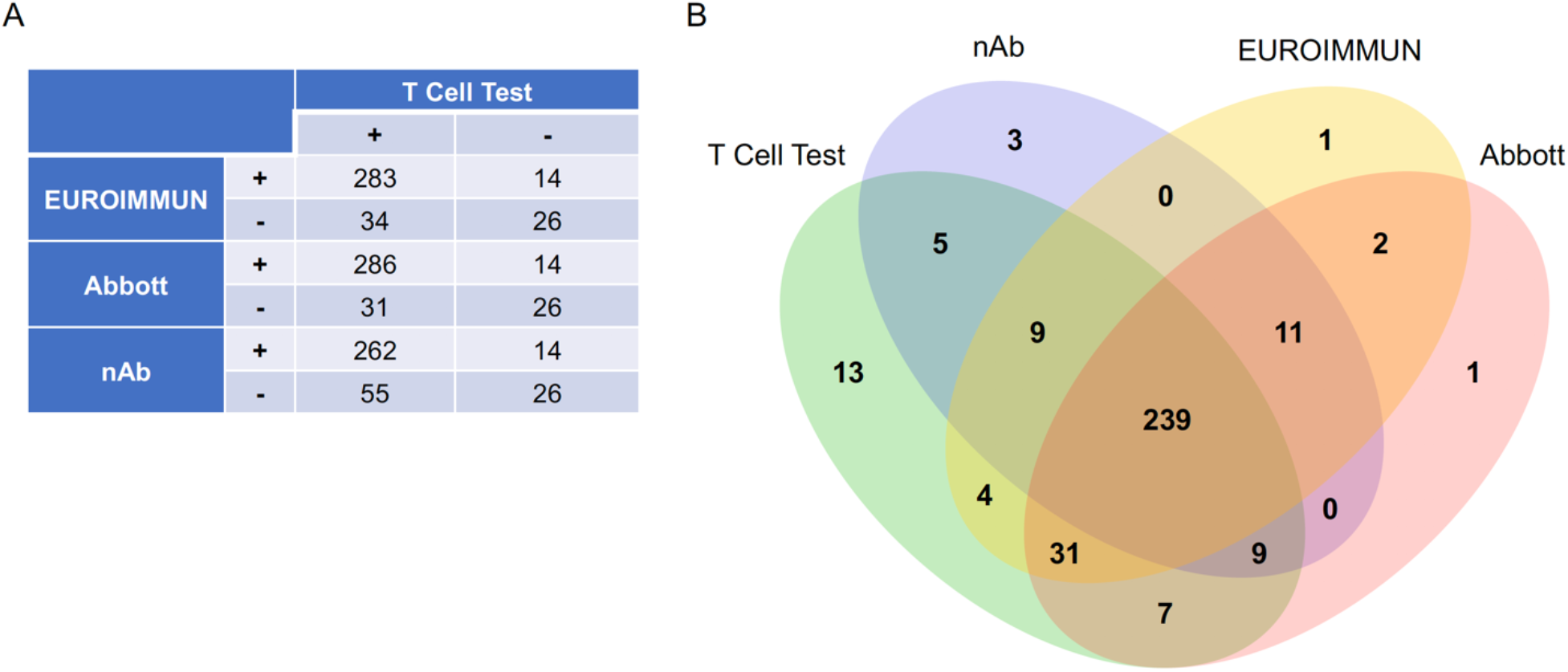
**(A) Table comparing the number of concordant and discordant SARS-CoV-2-positive test results by different assays. (B) Shared SARS-CoV-2-positive test results by different assays**.

**Supplemental Figure 4:**
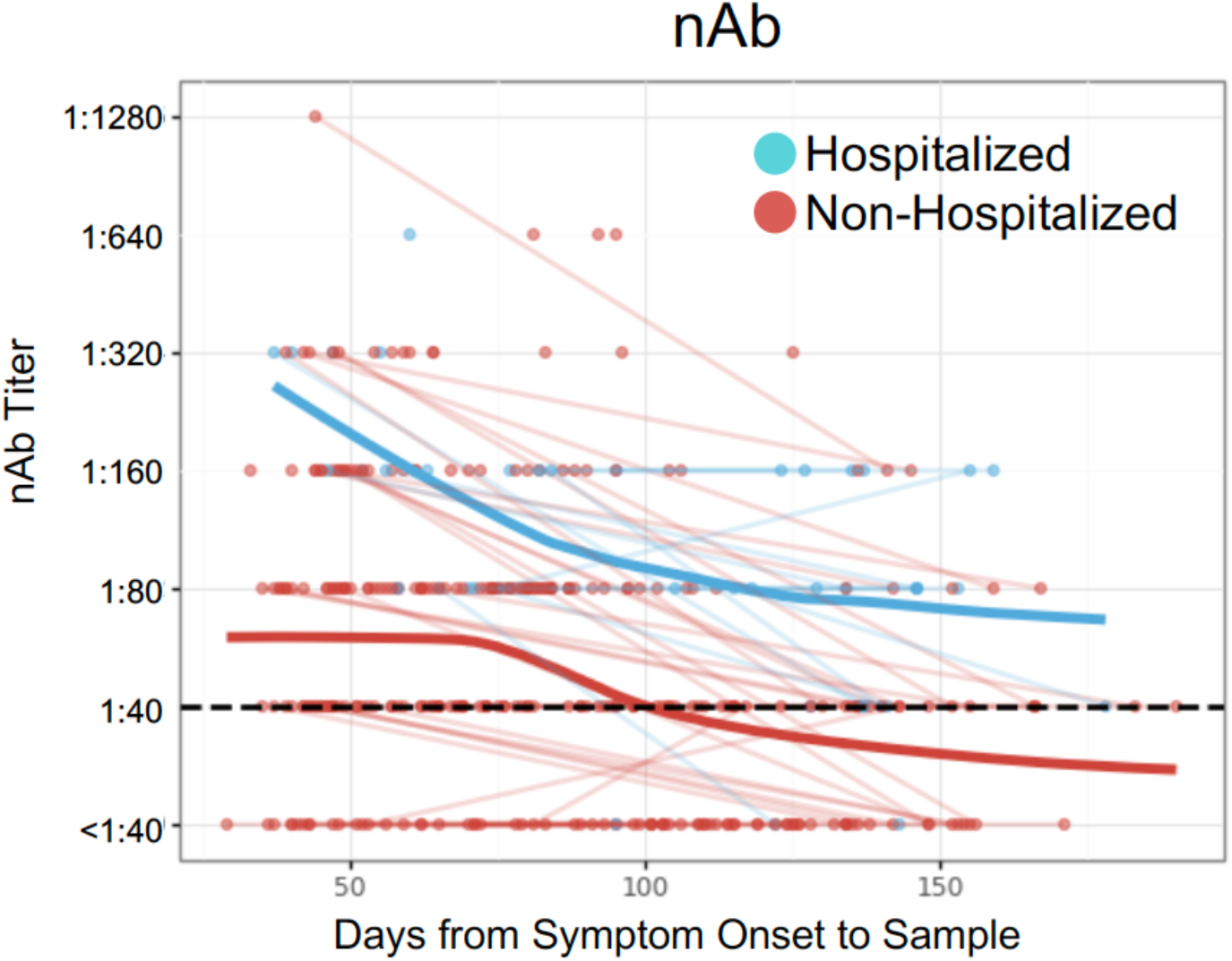
Neutralizing antibody titer (nAb) as a function of days from symptom onset to sample, indicated as hospitalized (blue) or non-hospitalized (red) individuals, with trend lines connecting visit 1 and visit 2 sample points from the same subject. Blue and red bold trend lines indicate smoothed mean (LOESS) for hospitalized and non-hospitalized individuals, respectively.

## Notes

### Author Declarations

Institutional Review Board approval was obtained from the University of Washington and all participants completed informed consent forms.

